# Probiotics treatment improves cognitive impairment in patients and animals: a systematic review and meta-analysis

**DOI:** 10.1101/2020.06.12.20129015

**Authors:** Tingting Lv, Mengfei Ye, Fangyi Luo, Baiqi Hu, Anzhe Wang, Jiaqi Chen, Junwei Yan, Ziyi He, Feng Chen, Chao Qian, Hui Gao, Jian Zhang, Zheng Liu, Zhinan Ding

## Abstract

The gut–brain axis has received considerable attention in recent years, and the “psychobiotics” concept indicates that probiotics have a potential positive effect on cognitive function. Therefore, the aim of this study was to quantitatively evaluate the influence of probiotics on cognitive function. We conducted a random-e?ects meta-analysis of 7 controlled clinical trials and 11 animals studies to evaluate the e?ects of probiotics on cognitive function. Probiotics supplementation enhanced cognitive function in both human (0.24 [0.05–0.42]; *I*^*2*^ = 0%) and animal studies (0.90 [0.47–1.34]; *I*^*2*^ = 74%). Subgroup analyses indicated that the effects of probiotics on cognitively impaired individuals (0.25 [0.05–0.45]; *I*^*2*^ = 0%) were greater than those on healthy ones (0.15 [−0.30 to 0.60]; *I*^*2*^ = 0%). Furthermore, compared with a multiple-probiotic supplement, one strain of probiotic was more effective in humans. The meta-analysis provided some suggestions for probiotics intervention and tended to support a customized approach for different individuals to ameliorate cognitive disorders. Future additional clinical trials are necessary to evaluate therapeutic effect and influencing factors.

## 1. Introduction

“Cognitive decline” as a common symptom refers to deterioration in cognitive ability to varying degrees(Langa et al., 2014). Cognitive decline is a growing public-health concern and very common in a variety of conditions, including aging, adiposity, depression, and especially in Alzheimer disease (AD) (Small, 2016). The World Health Organization (WHO) reports that about 50 million people now suffer dementia, and that with one new case every 3 s the number of dementia patients is set to triple by 2050 (WHO, 2019). The continuous increase will not only lead to a decline in quality of life but also contribute to profound social and financial consequences (Wortmann, 2012). However, pharmacological treatment is limited, and no drugs to completely reverse the symptoms are available; related treatments are not effective in all of the patients and can even generate undesirable side effects (Galimberti et al., 2010). Therefore, finding a therapy to promote cognitive function is urgent.

The gut microbiome has emerged in recent decades as a critical factor affecting neurophysiological and psychophysiological functions, including cognition, emotion neurotransmission, and neurodevelopment (Sarkar et al., 2018). The interaction between gut microbes and the central nervous system (CNS) exists within the so-called “gut–brain axis” (Rosshart et al., 2017), a complex bidirectional-association network between intrinsic gut microbes and the brain (Palm et al., 2015). In recent years, several lines of studies have suggested that changes in the composition of the intestinal microbiota (including in germ-free [GF] mice), such as those created by bacterial infections, antibiotics or supplements of probiotics, significantly influence cognitive function positively or negatively (Collins et al., 2012;Gareau, 2014;Gareau et al., 2011;Messaoudi et al., 2011). Probiotics are defined as living microörganisms with potential health benefits to the host when administered (de J R De-Paula et al., 2018); as one means of changing the composition and function of gut microbiota, probiotics have proven capable of affecting cognitive function (Sarkar et al., 2018). Therefore, our supposition was that probiotics supplementation might be an effective therapy for improving cognitive function through optimizing the composition and function of gut microbiota.

However, although some researchers report that probiotics supplements can improve cognition, relevant data are still scarce, and various studies have had inconsistent results(Bhattacharjee et al., 2013;Davari et al., 2013). Furthermore, whether probiotics promote cognition lacks conclusive results in both humans and animals, and multiple sources of discrepancies also exist. Up to now, no relevant meta-analysis has yielded a positive and consistent conclusion. Therefore, the objective of our meta-analysis was to provide quantitative results of probiotics’ effects on cognitive function in humans and animals, as well as provide some suggestions on how to use potentially suitable probiotics.

## 2. Methods

### 2.1 Literature search strategy

We conducted a systematic literature search to identify studies on the influence of probiotics supplements on cognitive function in humans and animals. We searched three electronic databases (Cochrane Library, PubMed, and EMBASE) to find potentially pertinent articles up through December 2019 using Medical Subject Headings (MeSH) and free-text terms for “probiotics” and “cognition” (Box 1). The study language was limited to English. We screened the returned titles to exclude duplicate studies and reviews, and then selected articles to exclude inappropriate and include appropriate titles and abstracts. Subsequently, we determined the final inclusion by reviewing the full text of the remaining studies. Also, we searched the references of included articles for additional studies to ensure that our search strategy discovered all relevant studies.

### 2.2 Inclusion and exclusion criteria

All of the studies included from the initial search strictly met the criteria of the Population, Intervention, Comparison, Outcome, and Study design (PICOS) framework according to Preferred Reporting Items for Systematic Reviews and Meta-Analyses (PRISMA) recommendations (Shamseer et al., 2015).

Humans: (1) Studies of adults age ≥18 years. Because of differing criteria for cognitive assessment between studies, there were no limitations on baseline cognitive status. (2) Studies that included pregnant women, or patients who had gastrointestinal disorders or had undergone gastrointestinal surgery, were excluded. (3) Experiments in probiotics intervention (any dose, strain, or administration method) to improve cognitive function were included. (4) We set the same conditions for the control group, except that it had no probiotics interventions. (5) At least one outcome of cognitive function was measured. (6) The study design was randomized controlled trial (RCT).

Animals: (1) Subjects were female and male rodents with or without cognitive impairments. (2) Intervention was probiotics (any dose, strain, or administration method) of at least 3 days’ duration. (3) Passive or active control was included in the study. (4) Outcomes of cognitive function were measured using the Morris water maze (MWM) test, Y-maze test, novel-object recognition test (ORT), and passive-avoidance test (PAT). (5) The study design was RCT. Studies without full text or primary data that could not be electronically extracted were excluded.

### 2.3 Data extraction

Two authors (TL and FL) read through each study independently to extract data and experimental details into a templated table that included the following information: author(s), publication year, country, population characteristics (e.g., age, gender, sample size, and sample type), probiotic characteristics (e.g., types and duration), and outcome measures. For animal studies, we also added species, model methods, and cognitive tests. When relevant data were available, we extracted the mean change, the SD of the mean change, and the number of participants in each group. If these data were not available, SE information was converted to SD, and the mean change was calculated according to baseline and post-intervention cognition scores. When specific data were not available, only figures or graphs, we requested unpublished data from authors. If we could not obtain these data, we used a digital ruler to estimate data from the figures or graphs (Song et al., 2017;Xie et al., 2014). Primary data were estimated based on the coordinate axis, and the mean and SD were calculated by statistical methods.

### 2.4 Risk-of-bias and quality assessment

Two authors independently assessed the risk of bias using the Cochrane collaboration tool (Higgins et al., 2011), judging the risk as “low,” “high,” or “unclear” in the following aspects: performance bias, selection bias, detection bias, reporting bias, attrition bias, and other biases. Uncertainties and discrepancies were resolved through discussion; when there was a disagreement, a third author became involved, and a consensus was reached.

### 2.5 Data synthesis and analysis

In the human studies, we measured the effects of probiotics intervention by the standardized mean difference (SMD) of the mean change from baseline. In the animal studies, due to the lack of baseline data, we calculated the effects of intervention using the SMD of the post-intervention cognitive variable between the intervention and control groups. Consistent with convention, the SMD was assessed using Hedges’ *g* as a measure of effect size; 0.2–0.5 was interpreted as small, 0.5–0.8 as medium, and ≥0.8 as large. When studies included ≥1 active-intervention group and multiple measure outcomes of cognitive variables, data of all comparisons were extracted and assessed. When cognitive outcomes were negatively scored (higher scores = decline in cognitive function), we reversed the computed outcomes of the effect size so that all of the positive scores reflected improvements in cognitive function. We used a random-effects model because we supposed that the effect sizes of included studies were comparable but not identical due to foreseeable heterogeneity across studies (e.g., different types of probiotics and measures of cognition) (Borenstein et al., 2010).

Subgroup analyses were conducted to assess whether effect size was affected by probiotic species, duration of regimen, or cognitive-outcome measures and to assess heterogeneity. We tested between-studies heterogeneity using Cochran’s Q test and *I*^*2*^ statistics. *I*^*2*^ values of 25%–50%, 50%–75%, and ≥75% respectively indicated low, moderate, and high heterogeneity (Higgins et al., 2003). To find possible sources of heterogeneity, we performed sensitivity analyses. Data were described as effect size ± 95% confidence intervals (CIs). We performed statistical analyses using Stata software version 12.0 (StataCorp, College Station, TX, USA). *P*-values < 0.05 were considered to be statistically significant.

## 3. Results

### 3.1 Study characteristics

The process of study selection is illustrated in Figure 1. Our initial search strategy identified 3451 records: 578 from PubMed, 170 from Cochrane, 2699 from EMBASE, and 4 from other sources. Duplicate records were removed and irrelevant studies eliminated by keyword (review, case, report, letter, and meta-analysis), after which we screened titles and abstracts. This left 205 articles to be assessed by reading the full text. We removed 187 of these articles because they had inappropriate experimental designs and interventions or lacked full text and related data, cognitive-outcome measures, or secondary analysis. Ultimately, 18 studies were included in the meta-analysis.

**Fig. 1.**
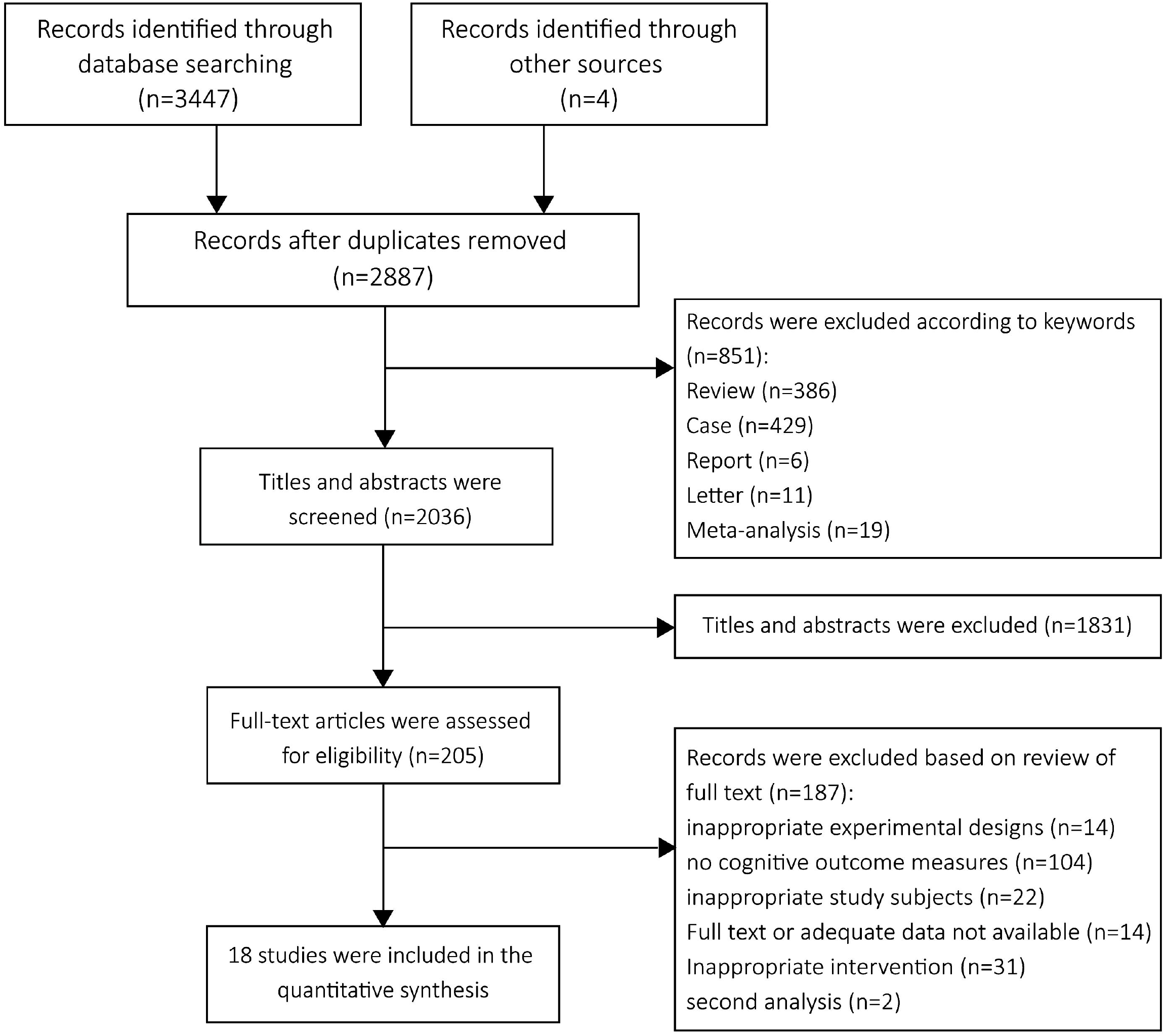
Selection process for trials included in the meta-analysis.

The general characteristics of these 18 studies are described in Tables 1 and 2. Seven were human studies (Agahi et al., 2018;Akbari et al., 2016;Bajaj et al., 2014;Inoue et al., 2018;Roman et al., 2018;Rudzki et al., 2019;Tamtaji et al., 2018) involving a total of 320 subjects, and 11 were animal studies (Avolio et al., 2019;Beilharz et al., 2018;Chunchai et al., 2018;Corpuz et al., 2018;Davari et al., 2013;Goudarzvand et al., 2016;Kobayashi et al., 2017;Liang et al., 2015;Luo et al., 2014;Nimgampalle et al., 2017;Tillmann et al., 2019)involving total of 318 subjects. Half of the 18 studies applied a single species of probiotic (n = 9), while the rest used ≥2 species of probiotics (n = 9). In three studies, in addition to probiotics/placebos, both intervention and control groups received an additional intervention: a trace element (selenium) (Tamtaji et al., 2018), moderate resistance training (Inoue et al., 2018), or selective serotonin reuptake inhibitors (SSRIs) (Rudzki et al., 2019). Study duration ranged from 8 days to 43 weeks, and daily doses of probiotics ranged between 1×10^8^ and 2.5 × 10^10^ colony-forming units (CFUs). All of the seven human studies included patients with AD (n = 3), fibromyalgia (FMS; n = 1), major depressive disorder (MDD; n = 1), minimal hepatic encephalopathy (MHE; n = 1) and healthy older adults (n = 1). Of the animal studies, two used mice, eight used rats, and one used hamsters. Cognitive outcomes were measured by the Y-maze test, MWM test, ORT, and PAT.

**Table 1.**
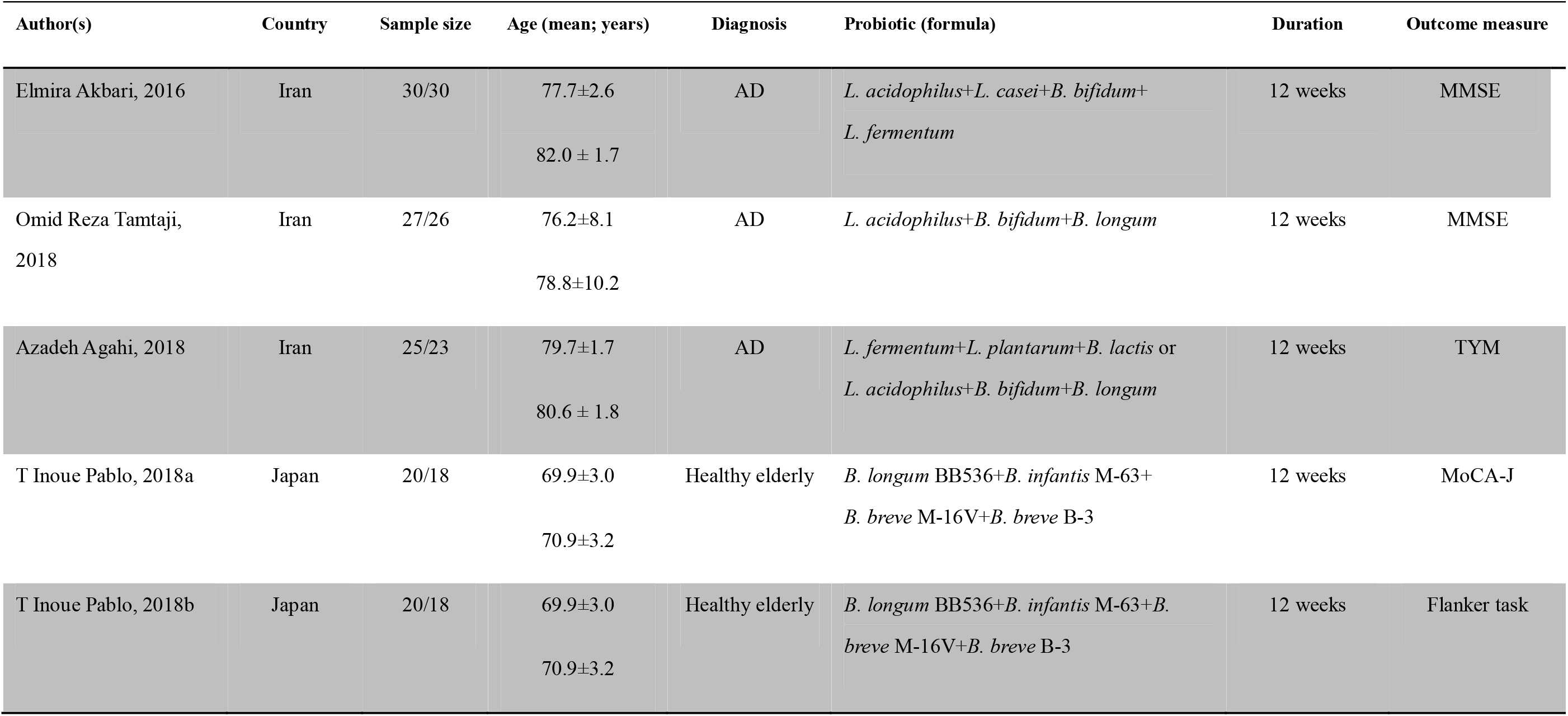
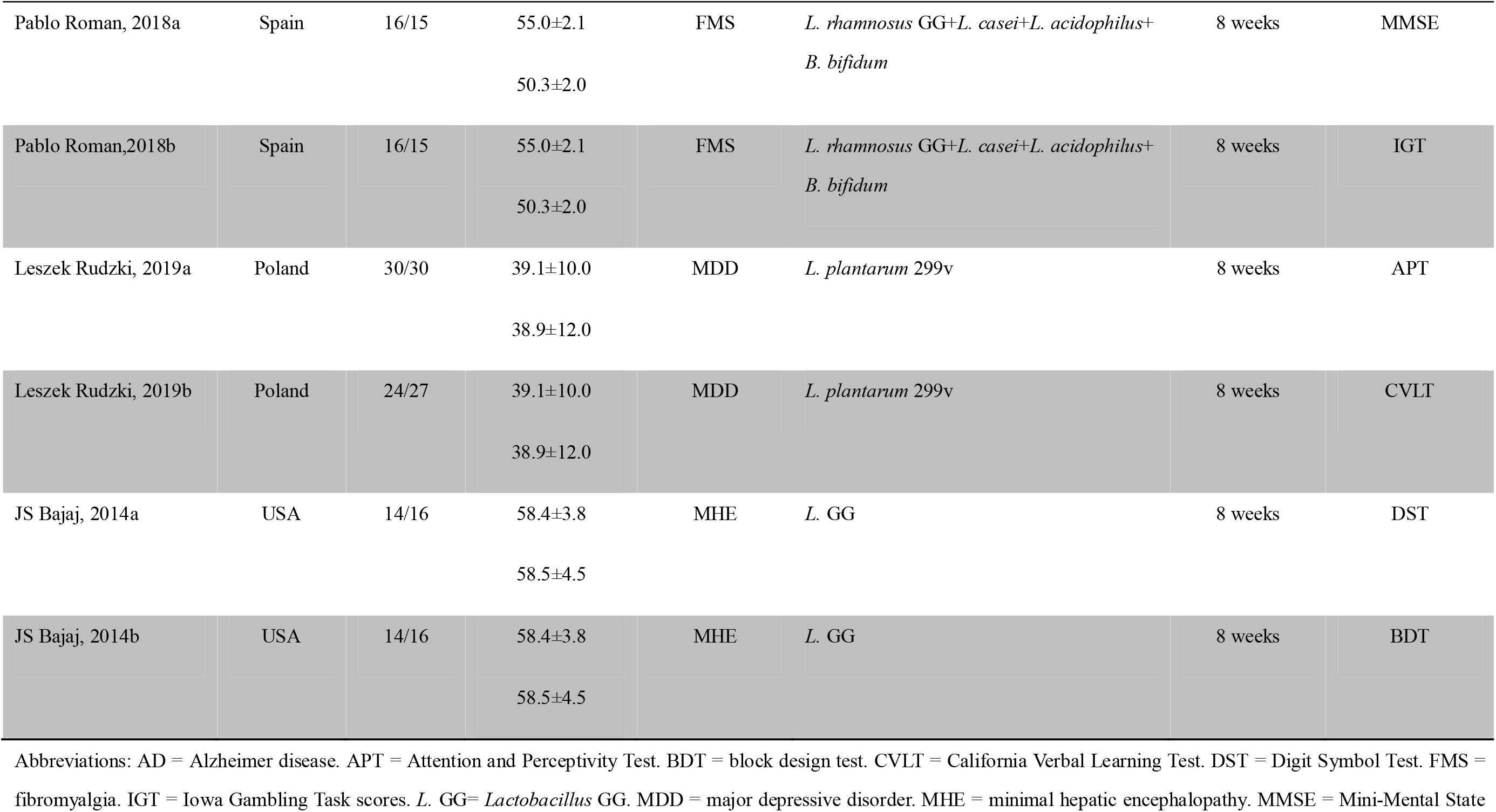
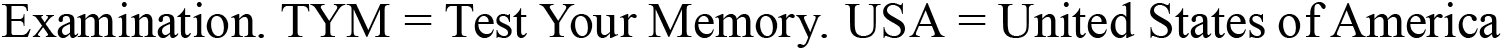
Characteristics of included studies in humans.

| Author(s) | Country | Sample size | Age (mean; years) | Diagnosis | Probiotic (formula) | Duration | Outcome measure |
| --- | --- | --- | --- | --- | --- | --- | --- |
| Elmira Akbari, 2016 | Iran | 30/30 | 77.7±2.6<br>82.0 ± 1.7 | AD | *L. acidophilus*+*L. casei*+*B. bifidum*+<br>*L. fermentum* | 12 weeks | MMSE |
| Omid Reza Tamtaji, 2018 | Iran | 27/26 | 76.2±8.1<br>78.8±10.2 | AD | *L. acidophilus*+*B. bifidum*+*B. longum* | 12 weeks | MMSE |
| Azadeh Agahi, 2018 | Iran | 25/23 | 79.7±1.7<br>80.6 ± 1.8 | AD | *L. fermentum*+*L. plantarum*+*B. lactis* or<br>*L. acidophilus*+*B. bifidum*+*B. longum* | 12 weeks | TYM |
| T Inoue Pablo, 2018a | Japan | 20/18 | 69.9±3.0<br>70.9±3.2 | Healthy elderly | *B. longum* BB536+*B. infantis* M-63+<br>*B. breve* M-16V+*B. breve* B-3 | 12 weeks | MoCA-J |
| T Inoue Pablo, 2018b | Japan | 20/18 | 69.9±3.0<br>70.9±3.2 | Healthy elderly | *B. longum* BB536+*B. infantis* M-63+*B. breve* M-16V+*B. breve* B-3 | 12 weeks | Flanker task |
| Pablo Roman, 2018a | Spain | 16/15 | 55.0±2.1<br>50.3±2.0 | FMS | *L. rhamnosus* GG+*L. casei*+*L. acidophilus*+<br>*B. bifidum* | 8 weeks | MMSE |
| Pablo Roman, 2018b | Spain | 16/15 | 55.0±2.1<br>50.3±2.0 | FMS | *L. rhamnosus* GG+*L. casei*+*L. acidophilus*+<br>*B. bifidum* | 8 weeks | IGT |
| Leszek Rudzki, 2019a | Poland | 30/30 | 39.1±10.0<br>38.9±12.0 | MDD | *L. plantarum* 299v | 8 weeks | APT |
| Leszek Rudzki, 2019b | Poland | 24/27 | 39.1±10.0<br>38.9±12.0 | MDD | *L. plantarum* 299v | 8 weeks | CVLT |
| JS Bajaj, 2014a | USA | 14/16 | 58.4±3.8<br>58.5±4.5 | MHE | *L. GG* | 8 weeks | DST |
| JS Bajaj, 2014b | USA | 14/16 | 58.4±3.8<br>58.5±4.5 | MHE | *L. GG* | 8 weeks | BDT |

Abbreviations: AD = Alzheimer disease. APT = Attention and Perceptivity Test. BDT = block design test. CVLT = California Verbal Learning Test. DST = Digit Symbol Test. FMS = fibromyalgia. IGT = Iowa Gambling Task scores. *L. GG*= *Lactobacillus* GG. MDD = major depressive disorder. MHE = minimal hepatic encephalopathy. MMSE = Mini-Mental State
Examination. TYM = Test Your Memory. USA = United States of America

**Table 2.**
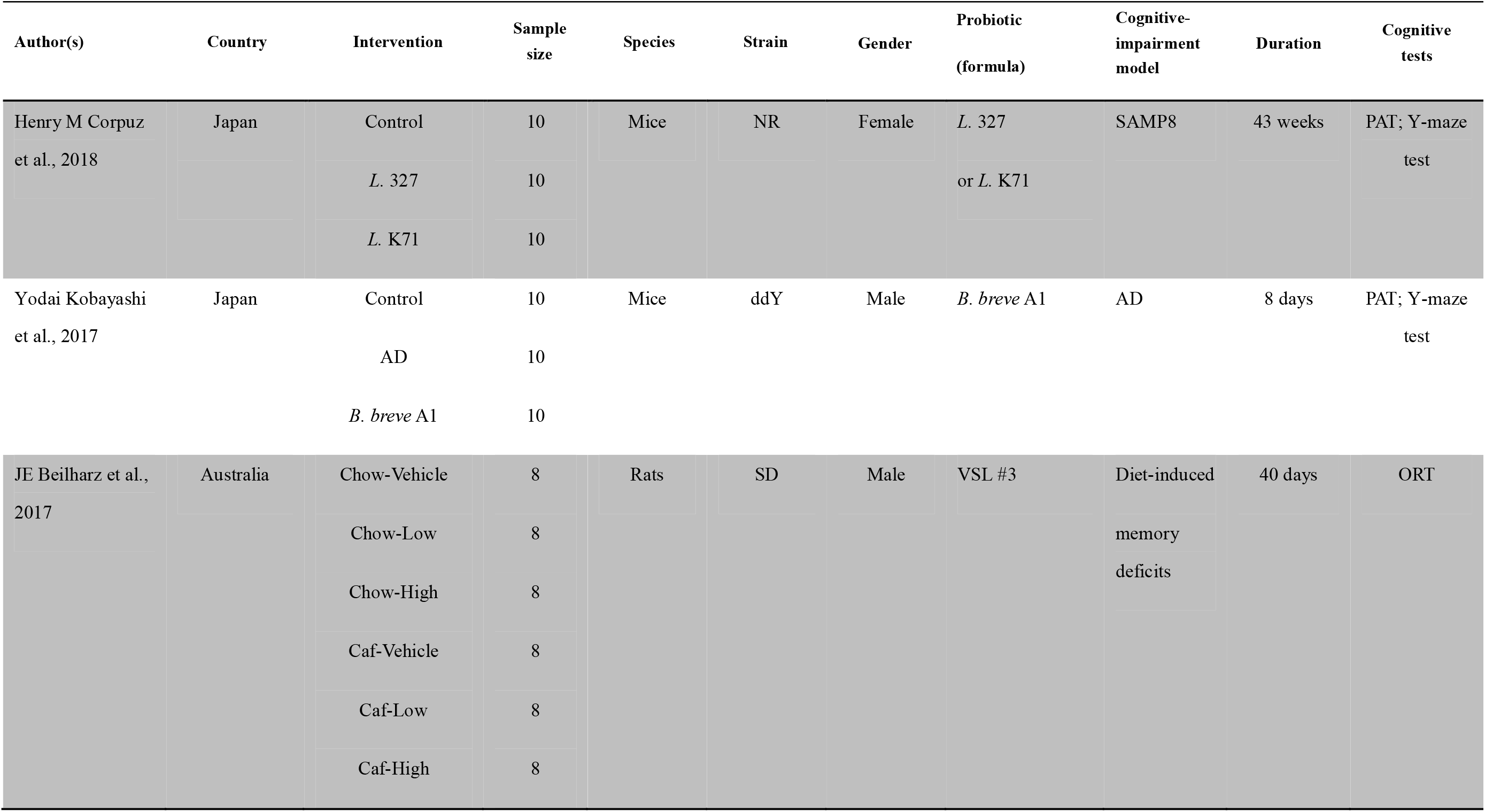
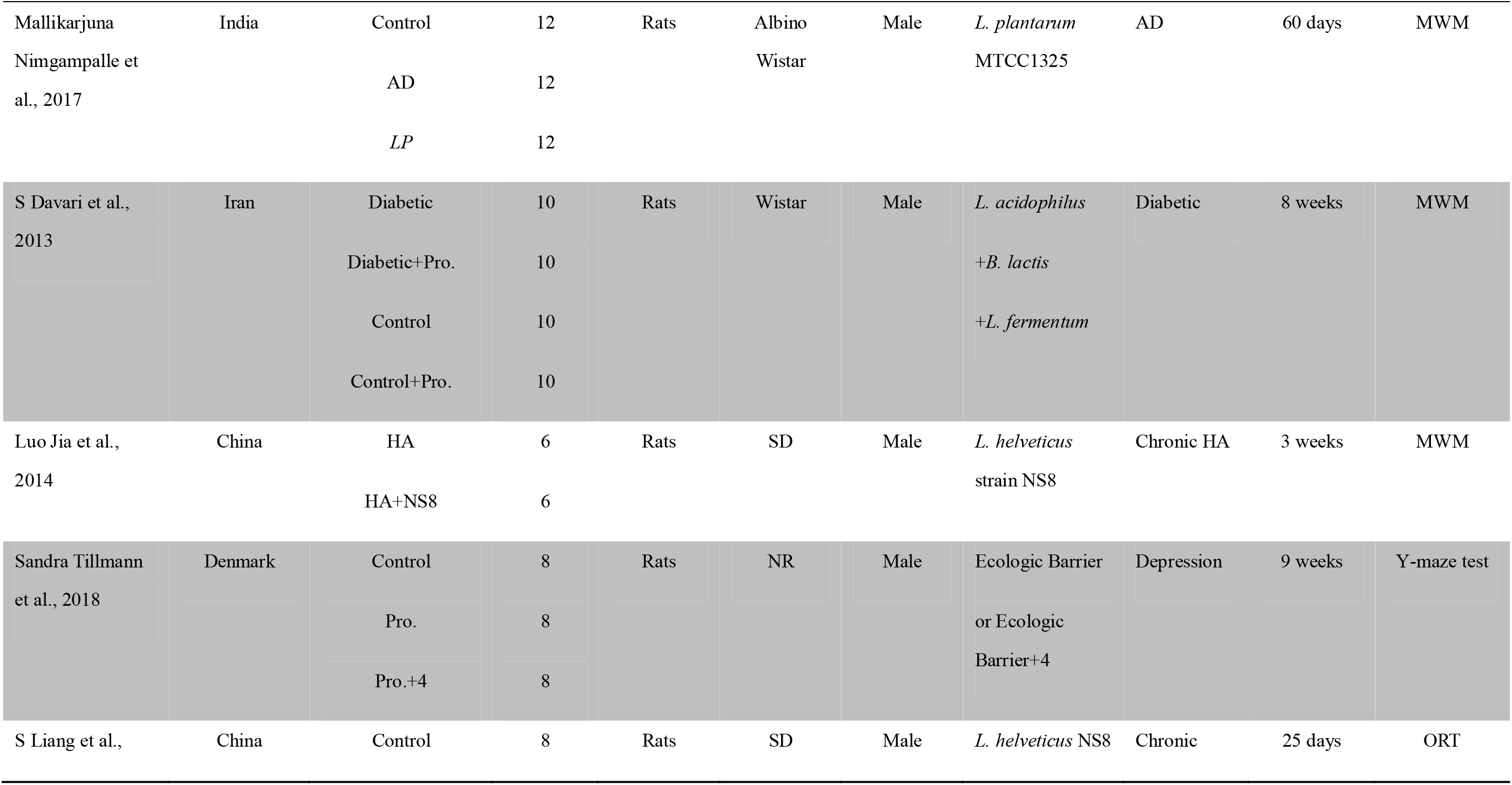
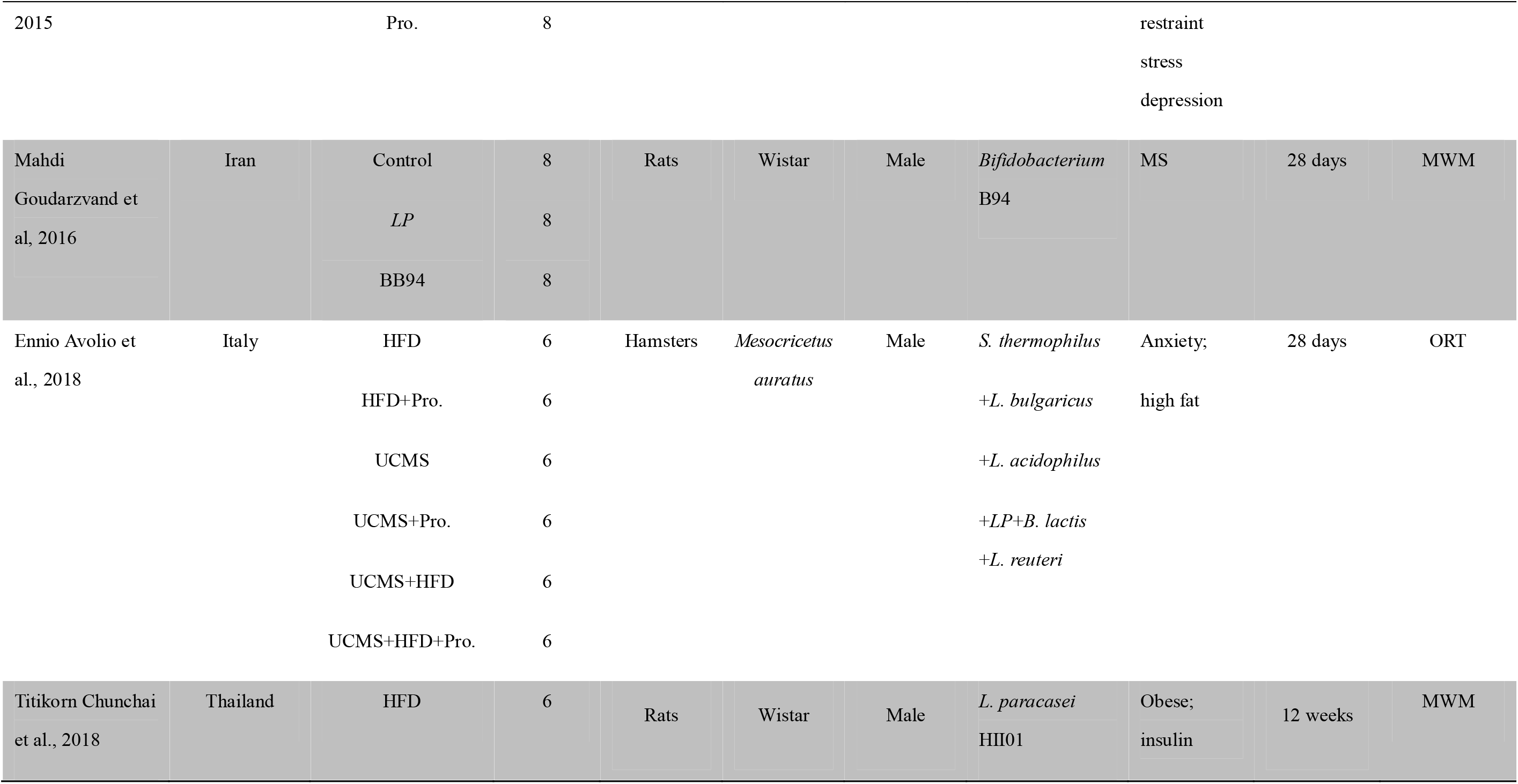
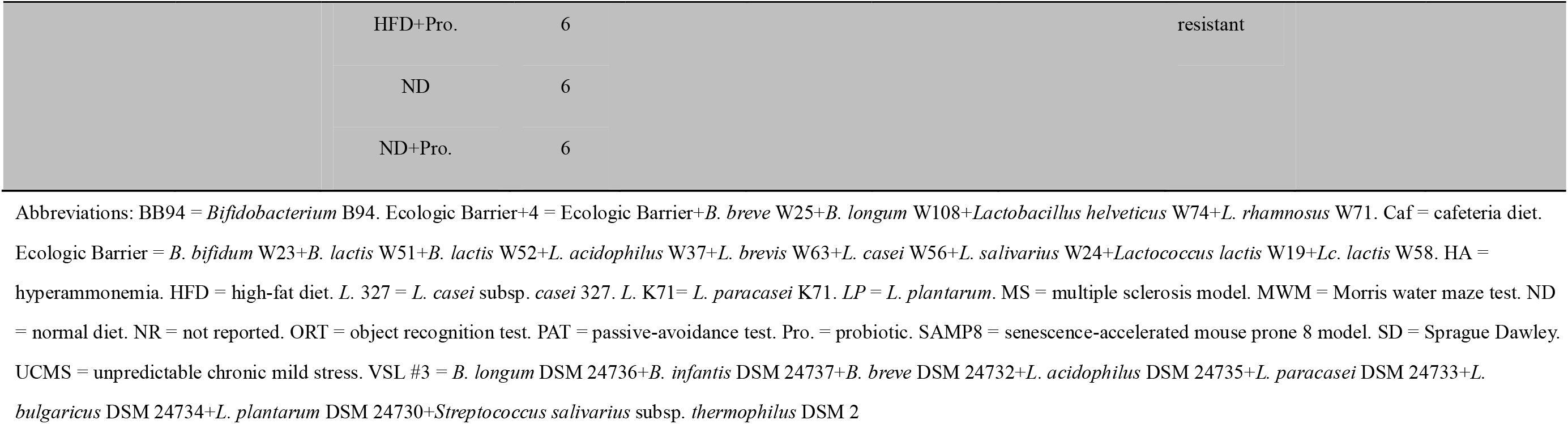
Characteristics of included studies in animals.

### 3.2 Study quality

#### Humans

All of the seven studies were randomized, and the majority (n = 5) described the methods of randomization. Most (n = 4) did not describe methods of allocation concealment in sufficient detail, but almost all (n = 6) were double blinded (participants and personnel). This indicated that the risk of selection bias was “high,” and that of performance bias was “low.” Although most of the studies did not mention detection bias, cognitive outcome was quantitatively measured by scales or various tests. Therefore, the assessor factor had little effect, and the authors believed that the risk of bias was “low.” Due to being unblinded, one study had a high risk of bias (Bajaj et al., 2014). Studies (n = 5) judged to have low risk of bias for incomplete outcome data included those using intention to treat principles in the data analysis, those providing reasons for exclusions, and those evenly distributing the number of dropouts between intervention and control groups. Studies regarded as having a high risk of other bias included those lacking *a priori* sample size analysis and those having images instead of concrete data (Fig. 2A).

**Fig. 2.**
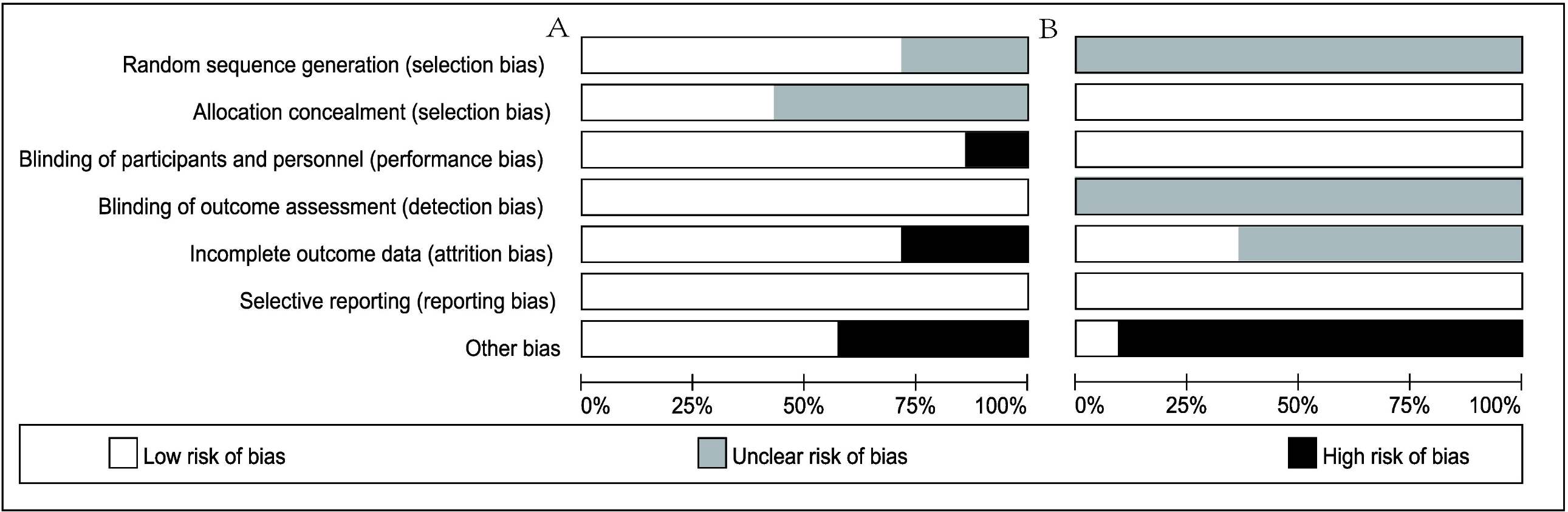
Risk-of-bias assessments of the included studies (domains from the *Cochrane Handbook for Systematic Reviews of Interventions*). A. Risk-of-bias assessments of the included human studies. B. Risk-of-bias assessments of the included animal studies.

#### Animals

We identified all of the studies as “unclear” in terms of random-sequence generation; although most reported randomization, no detailed methods of randomization were provided. Blindness and allocation concealment were not mentioned in all studies, but in animal studies they were assumed to involve “low” risk of bias. Risk of other bias was defined as “high” when articles had images instead of concrete data; since the data was acquired by software analysis, bias was inevitable. When relevant reference information was lacking, we defined the study as having “unclear” risk of bias (Fig. 2B).

### 3.3 Main efficacy of meta-analysis

#### Effect of probiotics on cognition

All of the seven human studies, involving 11 comparisons, reported cognitive outcomes. The summary effect size employing a random-effects model was 0.24 (95% CI, 0.05–0.42; *I*^*2*^ = 0%; *P* = 0.01; Fig. 3A). In the 11 animal studies, involving 25 comparisons, the overall data suggested that probiotics supplements were greatly effective in promoting cognition (0.90 [0.47–1.34]; *I*^*2*^ = 74%; *P* < 0.001; Fig. 3B), and moderate heterogeneity was present. We inferred that cognitive enhancement by probiotics in humans was inferior to that in animals.

**Fig. 3.**
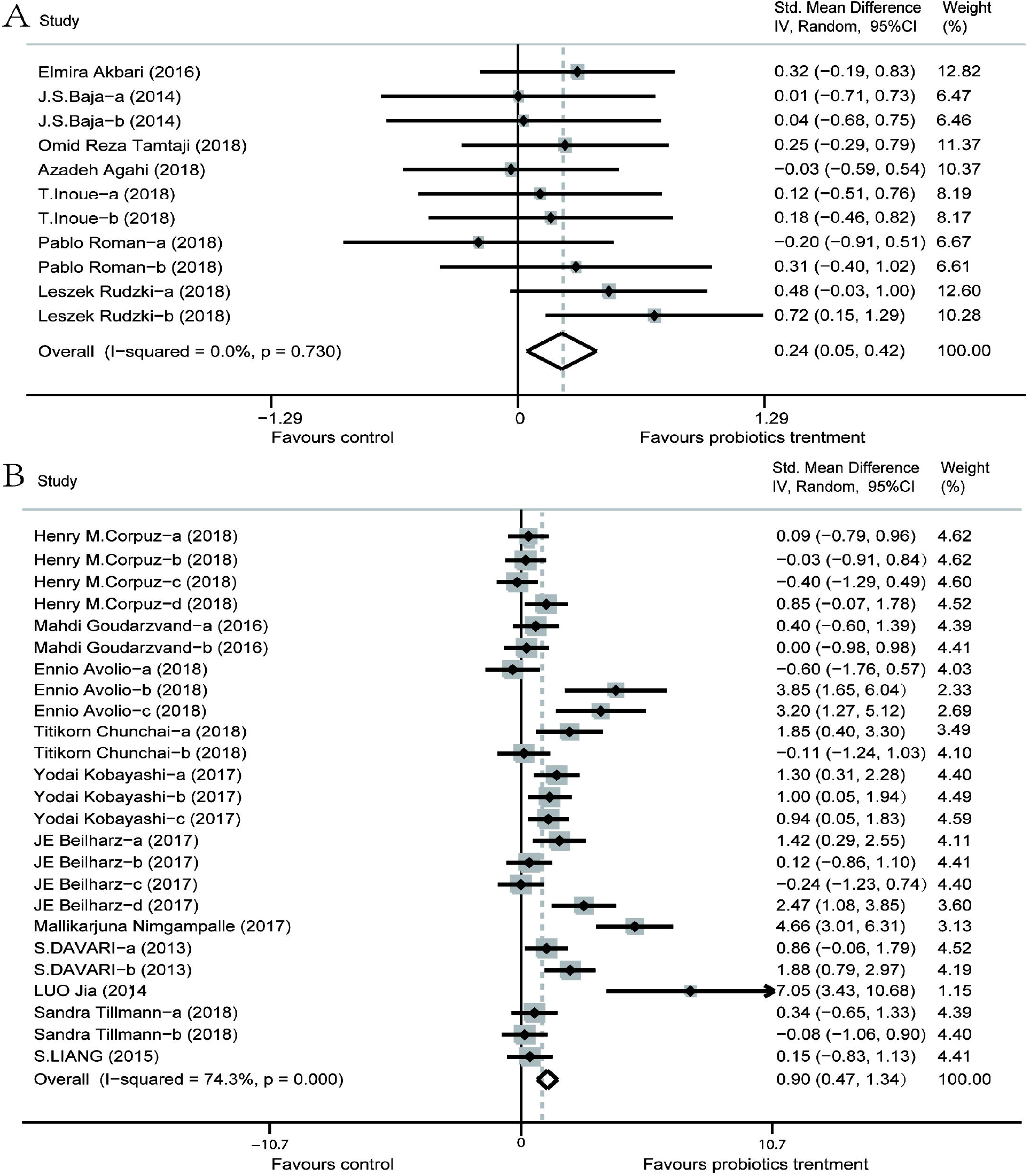
Forest plot of random-effects model meta-analysis of the overall effect of probiotics on cognition. A. The overall effect of the included human studies. B. The overall effect of the included animal studies.

#### Subgroup analyses

To investigate potential interfering factors in these results, we performed subgroup analyses of different variables at a time. The results of these subgroup analyses are summarized in Table S1 and described in detail below.

#### Experimental subjects

We conducted subgroup analyses of both human and animal studies. Based on whether subjects suffered cognitive impairment, we divided the studies into two subgroups. Our forest plot showed that for healthy individuals, probiotics intervention had no significant effect on promoting cognitive function either in humans (0.15 [−0.30 to 0.60]; *I*^*2*^ = 0%; *P* = 0.51) or in animals (0.40 [−0.54 to 1.34]; *I*^*2*^ = 69%; *P* = 0.40). For individuals with cognitive impairment, there was a small effect in human studies (0.25 [0.05–0.45]; *I*^*2*^ = 0%; *P* = 0.01) and a large effect in animal studies (1.02 [0.52–1.51]; *I*^*2*^ = 76%; *P* < 0.001; Fig. 4).

**Fig. 4.**
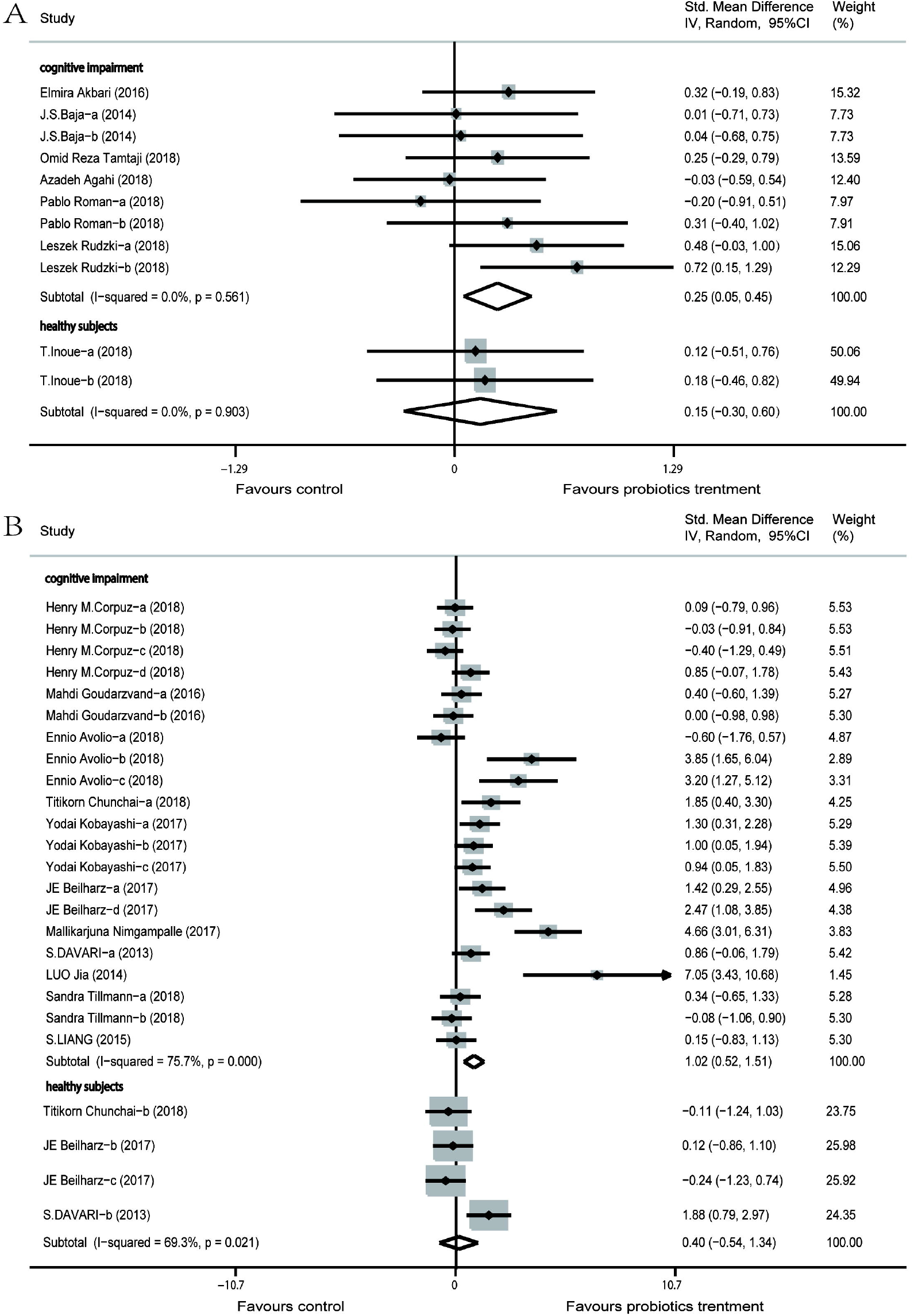
Forest plot of random-effects model subgroup analysis according to experimental subjects. A. Forest plot of the random-effects model subgroup analysis of human studies according to experimental subjects. B. Forest plot of the random-effects model subgroup analysis of animal studies according to experimental subjects.

#### Durations and strains of probiotics intervention

When we analyzed the data by probiotics intervention schedule (<12 weeks, ≥12 weeks), the forest plot indicated that a duration of <12 weeks only slightly enhanced cognition in human studies (0.29 [0.01–0.57]; *I*^*2*^ = 12%; *P* = 0.04) but greatly promoted it in animal studies (1.14 [0.59–1.68]; *I*^*2*^ = 77%; *P* < 0.001). Meanwhile, a duration of ≥12 weeks had no significant effect on cognition in either humans (0.18 [−0.08 to 0.44]; *I*^*2*^ = 0%; *P* = 0.17) or animals (0.27 [−0.29 to 0.82]; *I*^*2*^ = 46%; *P* = 0.34; Fig. 5).

**Fig. 5.**
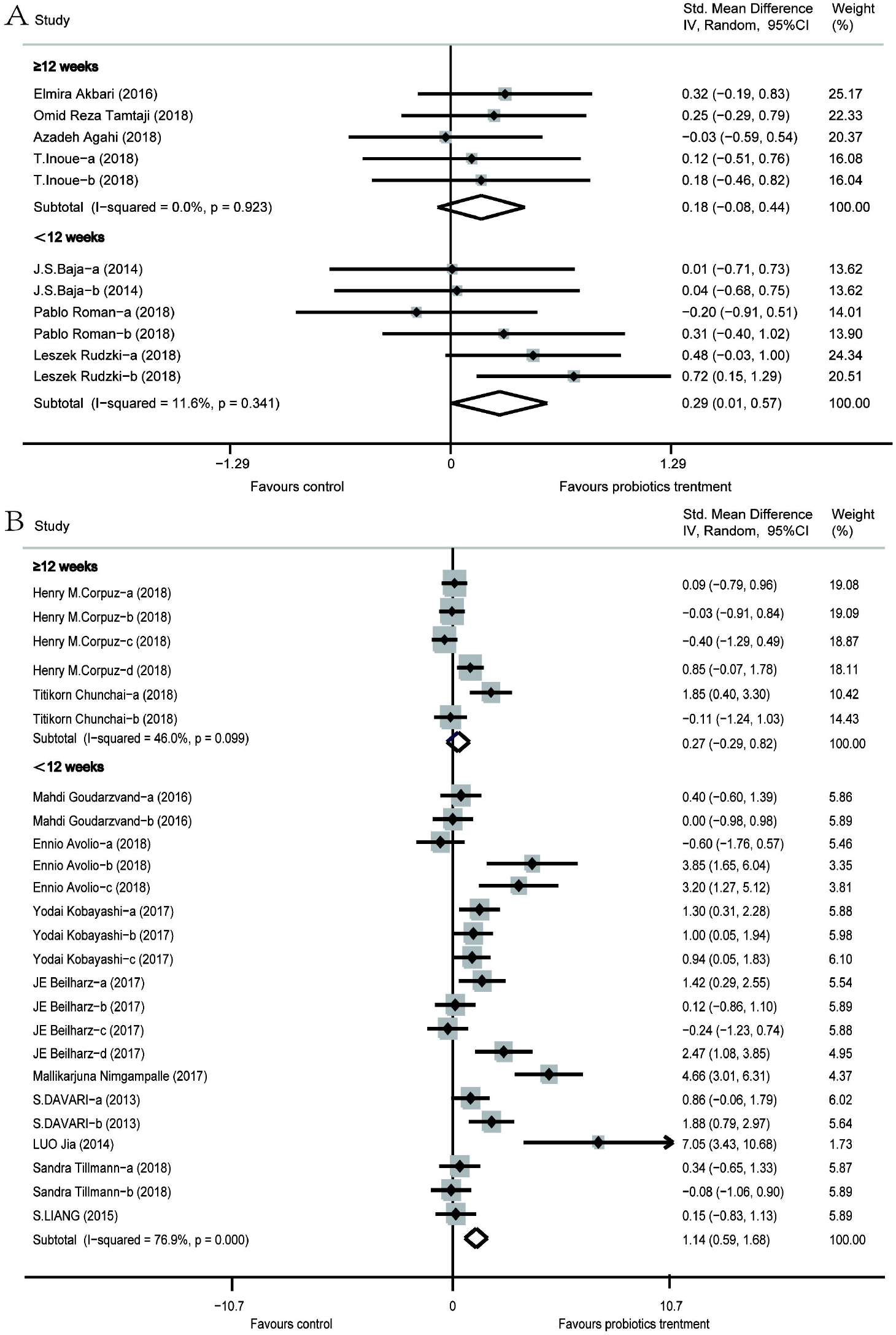
Forest plot of random-effects model subgroup analysis according to duration of probiotics intervention. A. Forest plot of the random-effects model subgroup analysis of human studies according to duration of probiotics intervention. B. Forest plot of the random-effects model subgroup analysis of animal studies according to duration of probiotics intervention.

Based on probiotics supplement formulations, subgroup analysis showed that when only one strain of probiotic was used, the effect on cognition was greater than when multiple strains were used. When supplied with one specific strain of probiotic, the forest plot showed a small effect in human studies (0.38 [0.05–0.71]; *I*^*2*^ = 13%; *P* = 0.02) and a large one in animal studies (0.84 [0.26–1.42]; *I*^*2*^ = 76%; *P* = 0.004). With regard to multiple probiotics supplements, the forest plot presented no significant effect in human studies (0.15 [−0.07 to 0.38]; *I*^*2*^ = 0%; *P* = 0.19) and a large effect in animal studies (0.99 [0.30–1.69]; *I*^*2*^ = 74%; *P* = 0.005; Fig. 6).

**Fig. 6.**
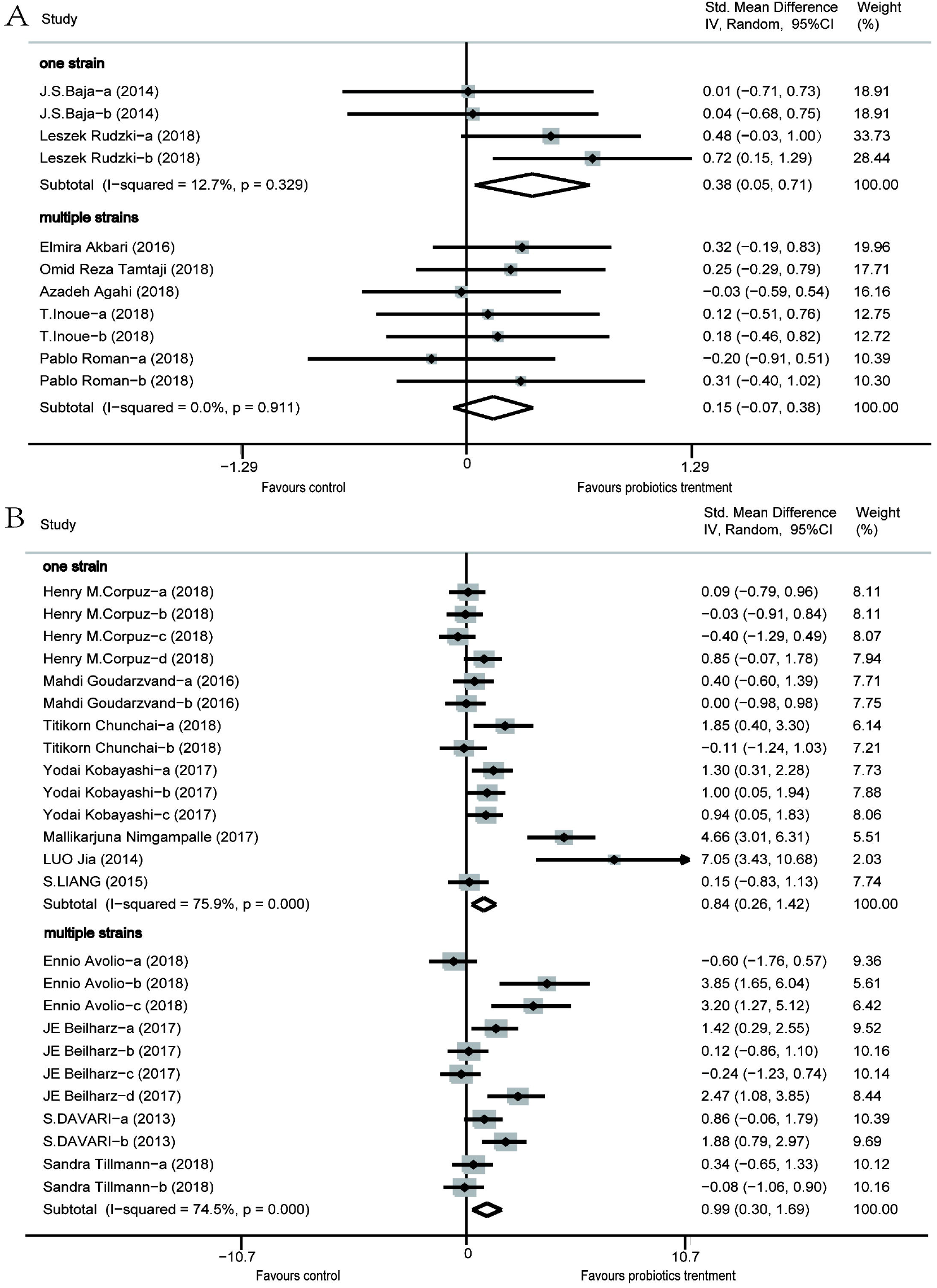
Forest plot of random-effects model subgroup analysis according to strains of probiotics supplement. A. Forest plot of the random-effects model subgroup analysis of human studies according to strain of probiotics supplement. B. Forest plot of the random-effects model subgroup analysis of animal studies according to strain of probiotics supplement.

#### Cognitive domains

We divided the human studies into two subgroups based on various cognitive domains. Global cognition was measured in one subgroup, and the result showed no significant effect (0.17 [−0.12 to 0.46]; *I*^*2*^ = 0%; *P* = 0.25). In the other subgroup, wherein various cognitive domains were measured by the corresponding tests, the results showed a small effect (0.28 [0.05–0.51]; *I*^*2*^ = 0%; *P* = 0.02). Animal studies were divided into four subgroups based on cognition tests. Those using the MWM test (−1.60 [−2.69 to −0.52]; *I*^*2*^ = 84%; *P* = 0.004) or the ORT (1.07 [0.12–2.02]; *I*^*2*^ = 79%; *P* = 0.03) to assess cognitive level indicated that probiotics greatly enhanced cognition, those using the Y-maze test showed a small effect (0.36 [−0.16 to 0.87]; *I*^2^ = 45%; *P* = 0.17), and those using the PAT showed a medium effect (0.59 [−0.06 to 1.23]; *I*^*2*^ = 33%; *P* = 0.07; Fig. 7).

**Fig. 7.**
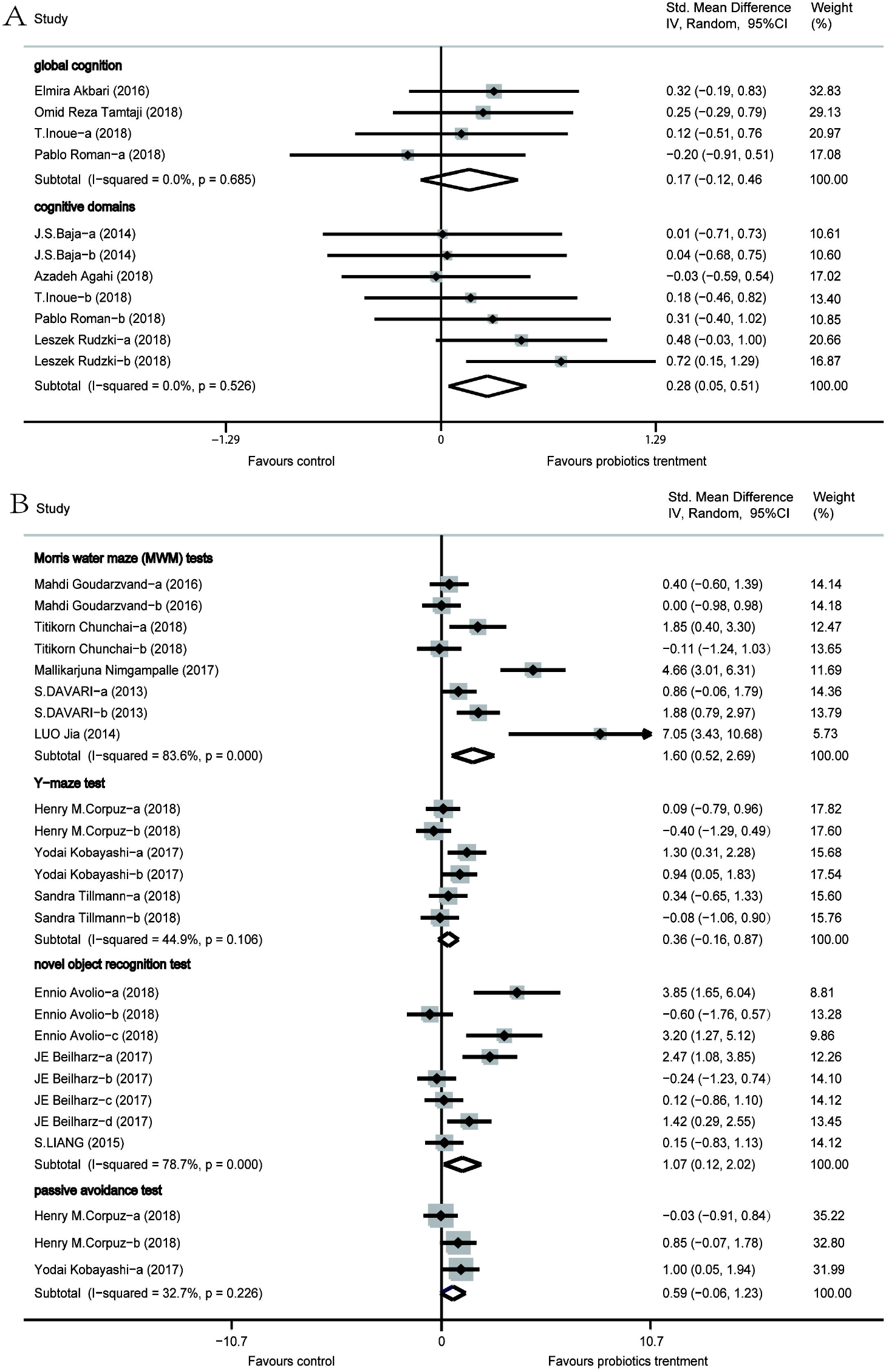
Forest plot of random-effects model subgroup analysis according to different cognitive domains. A. Forest plot of the random-effects model subgroup analysis of human studies according to different cognitive domains. B. Forest plot of the random-effects model subgroup analysis of animal studies according to different cognitive domains.

### 3.4 Heterogeneity analyses

Animal studies mainly demonstrated moderate heterogeneity. Via subgroup analysis, we identified the sources of heterogeneity, which could have been due to the cognition tests used, and the specific data are presented in Table S1. In subgroup analysis, the heterogeneity of the included animal studies decreased from 74% to 33%, indicating that the effect of probiotics supplements on cognition was correlated with different cognition tests.

## 4. Discussion

All of the human studies included were RCTs, which further enhanced the rigor and credibility of the studies. Based on our results, we found a positive overall effect of probiotics supplements on cognitive function. This effect was reflected in both humans (0.24 [0.05–0.42]; *I*^*2*^ = 0%; *P* = 0.01) and animals (0.90 [0.47–1.34]; *I*^*2*^ = 74%; *P* < 0.001). Furthermore, the included articles involved a variety of cognitive-disorder models, but the results still demonstrated positive effects on cognition. Therefore, we deduced that multiple forms of cognitive impairment, including aging, AD, diabetes, and depression, could be ameliorated by probiotics supplements via different mechanisms. Probiotics, as natural supplements, are well tolerated; no adverse events were reported, except that one probiotic presented a higher incidence of self-limited diarrhea (Bajaj et al., 2014). Accordingly, we deduced that probiotics would be an effective therapy for cognitive impairment.

Despite inadequate evidence that probiotics supplements ameliorate cognitive function, the connection between intestinal microbiota and neuropsychiatric function is well researched. The gut–brain axis partly optimizes bidirectional (top-down and bottom-up) regulatory interaction between the gut and the brain through endocrine, immune, and neurological pathways (Carabotti et al., 2015). The mechanism of probiotics supplements can be interpreted mainly through these three routes of communication. Evidence suggests that neuroinflammation mediates the deleterious effects on cognitive function of hyperammonemia, AD, and other diseases (Akbari et al., 2016;Rodrigo et al., 2010;Ryan et al., 2016). Probiotics are proven to prevent bacterial translocation, facilitate intestinal-barrier function, decrease inflammatory-cytokine levels, and suppress the activation of microglia, all of which lead to attenuation of systemic and neural inflammation (Gareau et al., 2007;Riedel et al., 2006;Zareie et al., 2006). Therefore, we hypothesized that one mechanism by which probiotics promote cognition was weakening neuroinflammation. The hippocampus plays a key role in cognition and could be a crucial mediator based on the above-described link between microbiota and cognition. Several studies have found that the use of probiotics significantly increased gene and protein expression of brain-derived neurotrophic factor (BDNF) and reduced oxidative stress and apoptosis in the hippocampus (Chunchai et al., 2018;Corpuz et al., 2018). BDNF is a critical neurotrophin that plays crucial roles in neuronal growth, survival, and plasticity, the last being important for cognitive processes (Yoshii et al., 2010). Neurotransmitter synthesis including gamma-aminobutyric acid (GABA), serotonin (5-HT), dopamine, acetylcholine, and norepinephrine might also be affected by probiotics (Cryan et al., 2012). Dysfunction of the 5-HT and GABAergic systems leads to memory and cognitive impairment (Castellani et al., 2008). Another mechanism might be that most *Lactobacillus* species and other probiotics excite the vagal nerves that connect with all of the neurons involved in behavioral changes such as depression, anxiety, learning, and memory (Wood, 2004). In terms of endocrine pathways, one study indicates that *L. helveticus* NS8 supplementation normalized most cognitive disorders by reducing corticosterone (CORT) release to modulate the function of the hypothalamic–pituitary–adrenal (HPA) axis and recovering BDNF, noradrenaline (NE), and 5-HT levels (Liang et al., 2015).

Our subgroup analysis of healthy subjects and patients indicated that in individuals with cognitive impairment, probiotic use improved cognitive function significantly more than in healthy individuals. Although only one human study used healthy subjects, we drew the same conclusion in our subgroup analysis of animal studies. It is easy to suppose that due to the ceiling effect, good baseline cognitive function might limit the scope for improvement. In the one study in healthy elderly subjects, improvement in cognitive function was observed after ingestion of probiotics, while no significant differences were found between the study’s two groups (Inoue et al., 2018). Similarly, in healthy rats, behavioral functions were not affected by probiotics administration (Davari et al., 2013). Therefore, we hypothesized that probiotics ameliorated cognitive focus in cognitively impaired individuals but not in healthy people. Even in one animal experiment, spatial memory was improved, but object memory declined after consumption of probiotics; the effect was obvious only in a later memory test, which meant that probiotics could be detrimental to healthy individuals and this effect might accumulate over time (Beilharz et al., 2018). We observed the same results in another study in humans (Benton et al., 2007). To the best of our knowledge, digestion of probiotics changes gut microbiota composition (de J R De-Paula et al., 2018). Therefore, we tend to support that probiotics might help restructure disordered gut microbiota toward health, whereas in healthy individuals this change might lead to abnormal composition or partially aberrant interaction pathways, depending on strain of probiotics or duration of use. Due to the scant number of studies with healthy subjects, we expect to explore additional related literature and more-detailed mechanisms in the near future.

Gut microörganism diversity might not always be beneficial (Sarkar et al., 2018). One study of 1-year-old infants proved that higher diversity was linked to poorer scores on the expressive-language scores of the Mullen Scales, Early Learning Composite (ELC), and visual-reception tests(Carlson et al., 2018). Meanwhile, high gut microörganism diversity has been found in adults with autistic-spectrum disorders and MDDs (Finegold et al., 2010;Jiang et al., 2015). Subgroup analysis of the number of probiotics used in human studies further supported this finding: when various probiotics supplements were used, the effect on cognitive function surpassed that yielded by a single strain of probiotic. Our corresponding subgroup analysis of animal studies did not reach the same result. On one hand, relatively high heterogeneity limited accuracy of the result; on the other, animal models have narrow translatability to humans. More types of probiotics do not mean more benefits to neurocognitive or neuropsychiatric function. Higher gut microörganism diversity could mean fewer resources afforded to each gut microörganism.

With regard to duration, subgroup analysis results showed that when probiotics intervention was <12 weeks, the beneficial effect on cognitive function was more obvious, in contrast to the conclusions of many studies that longer duration means better results. In the human studies, all of the AD patients took probiotics for 12 weeks; most of them were in a relatively severe condition, so enhancement of cognitive function was limited. Meantime, poor timing of intervention could influence effectiveness; in the later stages of AD, the development of neurofibrillary tangles means irreversible pathological changes (Brewer, 2010). Therefore, advance intervention might be more effective. In animal studies, only two trials ran for ≥12 weeks, which impaired extrapolation. Therefore, more-rigorous studies are needed to ascertain the effect of intervention duration on cognitive function.

Probiotic strains of *Lactobacillus* and *Bifidobacterium* were employed in most included studies, except for one that used strains of *Streptococcus* (Avolio et al., 2019). Many strains of *Bifidobacterium* and *Lactobacillus* have already been marketed in products for human consumption (Fujimoto et al., 2008). In many rodent models and human studies, strains of both these genera were found to improve memory and decrease the rates of depression- and anxiety-like behaviors (Ait-Belgnaoui et al., 2014;Castellani et al., 2008;Liu et al., 2019;Tillisch et al., 2013). One study confirmed that *L. helveticus* NS8, by enhancing indoleamine 2,3-dioxygenase (IDO) activity, modulates the kynurenine pathway to promote cognitive decline (Luo et al., 2014). Conversely, in a rat model of depression, treatment with *L. johnsonii* or *Bifidobacterium* spp. inhibited IDO production (Desbonnet et al., 2008;Valladares et al., 2013). This reflects that different strains lead to diverse results, and in our study we further assumed that different probiotics would ameliorate cognitive function via different mechanisms. It is difficult to suggest a particular strain of probiotic in this meta-analysis because inter-study discrepancies in dosing and duration of intervention impaired the comparability of included trials.

Although the overall effect in animal studies was significant, the moderate heterogeneity of these studies was also of concern. Therefore, we employed the most significant subgroup analysis (cognitive tests) to examine the sources of heterogeneity. Through such analysis, we found that probiotics supplements were greatly effective in promoting cognitive function, as demonstrated by MWM tests (−1.60 [−2.69 to −0.52]; *I*^*2*^ = 84%; *P* = 0.004) and ORTs (1.07 [0.12–2.02]; *I*^*2*^ = 79%; *P* = 0.03), and that these results were accompanied by high heterogeneity. Therefore, we inferred that the moderate heterogeneity of the overall effect size originated from cognitive tests. The high heterogeneity could be related to the following: (1) Animals belonged to different species (e.g., rats and hamsters) and different strains (Wistar and Sprague Dawley mice) with discrepant genetic and biological characteristics. This would have led to adaptation in cognitive tests, and so animals differed in their sensitivities to probiotics. Furthermore, we doubt that the cognitive-impairment models were more likely to determine probiotics’ effects on cognition; more research is needed to verify this. (2) The MWM test and ORT require animals to be in a relatively high state of stability, which is hard to absolutely guarantee. Therefore, we expect more-accurate, more-comprehensive experiments to be performed. In light of this subgroup analysis, meanwhile, we found that probiotics administration could significantly promote object memory and spatial learning and memory in animals. Regarding the subgroups in human studies based on cognitive domains, we found that when we measured global cognition by scales, the effects on cognition were less significant than when we measured cognitive domains by corresponding tests. We suspect that cognitive tests are more sensitive to cognitive change.

Animal research lays the groundwork for human research. It has significant advantages in establishment of models, control of experimental conditions, and accessibility of mechanism research. In addition, the results of animal studies can provide reference and enlightenment for human studies on interventional mechanisms and effects. However, because of significant differences in physical structure and psychology between animals and humans, the limitations of extrapolation in animal researches also should be considered. Therefore, their results are not always coincident. Meanwhile, various interference factors in human research and the unique social attributes of humans can also lead to different results between human and animal studies.

The major strength of our study is that we acquired cognitive data from eligible articles and reached a conclusive conclusion. In addition, this is the first meta-analysis to quantify the cognitive benefits of probiotics. Furthermore, the assessment of study characteristics by subgroup analysis provided insights into some detailed probiotics supplementation proposals. Despite this, several limitations of this meta-analysis must be considered. First, we acquired data using digital software, meaning there was a certain degree of data error. Second, we included participants with different types of diseases, and the studies used different models of cognitive impairment and examined different cognitive domains. However, subgroup analysis did not reveal any discrepancies in the effects of probiotics in different cognitive-impairment models. The differing effects are hard to deduce because there were different types, doses, and durations of probiotics supplementation. Third, inter-study differences in dosing, duration, and type made the results less convincing. Fourth, the amount of human research was limited. Fifth, evaluation of cognitive function lacked consistency and objectivity, which needed to be assessed via more-precise methods. Therefore, more high-quality studies are needed to confirm ideal probiotics strains/species, doses, and intervention durations.

## 5. Conclusion

Our meta-analysis showed that probiotics supplementation had an overall significant effect in promoting cognitive function in both humans and animals, and that probiotics could be effective and accessible cognitive therapy. Subgroup analyses found that probiotics supplements improved cognitive function in cognitively impaired individuals while having no significant effect on healthy people and possibly even being detrimental to them. Due to strain specificity and discrepancies in cognition-impairing conditions in subjects, we tend to support a customized approach for different individuals to ameliorate cognitive disorders toward a healthy state. Further high-quality studies should be conducted to learn the detailed mechanisms of various probiotics and to recommend formulations.

## Data Availability

We have confirmed.

## ACKNOWLEDGMENTS

This work was funded by the National Natural Science Foundation (nos. 81560059 and 81760058), Zhejiang Medical Health Science and Technology Project (no. 2019328893), the Scientific Research Fund of Shaoxing University (no. 20125025), and the National Training Program of Innovation and Entrepreneurship for College Students (no. 2017R10349001).

